# Modeling Biomarker-Guided Avoidance of Radical Cystectomy: Costs and Outcomes

**DOI:** 10.64898/2026.08.17.26360582

**Authors:** Tamir Sholklapper, Mengmeng Li, Abhishek Srivastava, Aditi Wagh, Elizabeth Handorf, J. Robert Beck, Philip H. Abbosh

## Abstract

**Importance:** There is growing interest to avoid radical cystectomy (**RC**) in patients with muscle-invasive bladder cancer (**MIBC**) who receive neoadjuvant chemotherapy and achieve pathological complete response (**ypCR**). To achieve this goal, molecular biomarkers will likely need to be used to enhance clinical staging given the limitations of evaluation by cystoscopy, cytology, and cross-sectional imaging. There are no studies evaluating whether safe RC avoidance (**SaRCA**) would be cost effective and what impacts it would have on quality of life (**QoL**) and survival.

**Objective:** This study models the potential economic, QoL, and survival costs/benefits of a ypCR biomarker as it relates to SaRCA using a decision analysis and Markov Model (**MM**).

**Methods/Materials:** A decision tree and MM was created to compare the expected costs of initial treatment, QoL, and survival under one strategy where all patients undergo RC after neoadjuvant treatment versus an alternative strategy where all patients would be subjected to the biomarker test with biomarker-positive patients (those with presumed residual disease) undergoing RC, while biomarker-negative patients (presumed complete responders) would undergo surveillance for up to 20 years. ypCR rates to neoadjuvant therapy, survival with and without RC, quality adjusted life years (**QALY**), and costs were abstracted from the literature. Test cost, sensitivity, and specificity were also abstracted from the literature for multiple clinical or liquid biopsy approaches.

**Results:** Broadly, SaRCA approaches are cost effective with the exception of systematic endoscopic evaluation (**SEE**). All testing approaches result in higher QALY and overall life expectancy compared to no testing. The cost of the test is offset by decreased usage of RC to realize a cost savings. These domains are further improved when cisplatin-based chemotherapy is replaced with emerging neoadjuvant therapies.

**Conclusions and relevance:** Modeling supports the development of accurate biomarker tests which can distinguish residual disease states to enable SaRCA. Such a biomarker could be used to avoid an expensive and risky operation, and unexpectedly would provide a survival benefit by reducing the number of perioperative mortalities in patients achieving ypCR. Development of an accurate biomarker-based test is likely to reduce cost and increase QoL and survival. An accurate biomarker test would have utility for patients, payers, hospitals, and physicians.

## Introduction

Radical cystectomy (RC) is a complicated procedure, with major complication rates of 15-30% and perioperative mortality rates similar to or exceeding that of open heart surgery in most published series^1–7^. Further, the cost of bladder cancer care is highest of all cancers per patient^8^ with RC incurring a $150K cost per patient in the first year after surgery - doubled if radiotherapy is used instead of RC^9^. RC also requires urinary diversion, a life-changing procedure.

Neoadjuvant chemotherapy (**NAC**) followed by RC is a standard of care treatment strategy for non-metastatic muscle-invasive bladder cancer (**MIBC**). Neoadjuvant cisplatin-based chemotherapy or immune checkpoint inhibitors have demonstrated pathological complete response (**ypCR**) rates ranging from 31-45%^10–14^. These results, along with the high complication rates, cost, and impacts on patient quality of life, have encouraged the exploration of RC avoidance in patients achieving *clinical* complete response (**ycCR**) after neoadjuvant therapies.

Several retrospective studies have evaluated patients who did not undergo RC after they were deemed to have achieved ycCR, defined as no evidence of tumor on cystoscopy/biopsy, exam under anesthesia, and cross sectional-imaging. This approach fails 30-50% of the time, where failure is defined as recurrence, need for salvage RC, metastatic disease, or death from bladder cancer^15–18^. Failure may be caused by occult (unrecognized) disease at the time of clinical restaging, which occurs in >50% patients in the only prospective trial evaluating undergoing such staging^19^. Further underscoring the poor performance of clinical restaging, 50% of patients deemed to have ahieved cCR or ycCR actually harbored MIBC at the time of RC. In contrast, retrospective studies show that up to 70% of patients who achieve ycCR avoid RC and have a long bladder-intact survival, sometimes requiring additional endoscopic resections^15–18^. Patients who achieve ypCR after NAC+RC achieve 85% long-term disease-free survival^20^, further suggesting that RC might be safely avoided in properly selected patients because it is rare for these patients to develop distant disease.

Published studies have shown a cost benefit for the use of NAC^21,22^ as well as cost benefit for *predictive* biomarkers of response^23^, however, cost effectiveness studies evaluating *prognostic* biomarkers, specifically for the purpose of RC avoidance, are lacking. Imaging tests such as pelvic MRI to detect residual disease have been developed for downstaging (ypT≤1) but appear to be much less accurate for assessing true ypCR^24^. Urine biomarkers developed for the purpose of prospective identification of patients achieving ypCR^25–27^ or to identify patients at risk for recurrence of nonMIBC^28–31^ have encouraging performance characteristics, however, blood or tissue tests could perform equally well. Our study to determine cost effectiveness is test-agnostic.

Clinical utility of a complete response test could be demonstrated if it either (1) improves QoL of patients undergoing the test, (2) increases overall survival in patients undergoing the test, or (3) reduces overall cost of treatment of MIBC. We used a Markov Model (**MM**) to estimate costs, survival, and QoL of MIBC patients undergoing either traditional RC or SaRCA (which we define as biomarker-informed RC avoidance) after cisplatin-based NAC or enfortumab-vedotin + pembrolizumab (**EVP**) and report the results herein.

## Methods

### Model inputs

A MM assumes that individuals within a large population can be assigned to a unique state with definable costs and QoL. Individuals can transition from one Markov state to another with defined choice-chance constants over time. By iteratively running the model using anticipated clinical outcomes and subsequent treatment algorithms, costs and impacts on life expectancy and QoL can be derived for comparison of multiple clinical approaches (i.e. a decision analysis). We used published literature or Center for Medicare/Medicaid Services tables to estimate (1) <u>costs</u> associated RC and its complications, surveillance procedures in patients undergoing SaRCA, treatment of localized or distant recurrences, using different SaRCA testing modalities; (2) <u>probabilities</u> of ypCR, perioperative mortality, survival after treatment; (3) choice-chance transitions from one Markov state to another; and (4) <u>QoL</u> of patients after bladder cancer treatment. When using Medicare/Medicaid costs, we used the 12502 MAC locality (Philadelphia, PA). The statistical performance of the SaRCA test was varied accordingly to evaluate the potential for cost effectiveness and changes in life expectancy and QoL. The tests chosen were MRI^24^, ctDNA^32^, SEE^33^, and two urine-based liquid biopsy tests^25–27^ and their sensitivities and specificities are described in each cognate reference as documented in Supplemental Table 4. For MRI, VI-RADS score ≤3 was considered to denote ycCR. Positive SEE, ctDNA or urine tests were considered to denote residual disease.

### Model description

Figure 1 depicts the decision trees used for this study. After NAC, the decision is to move to RC or perform biomarker testing and act on the results. If the biomarker is positive for residual disease (**RD**), RC is performed. If negative, the patient is placed on active surveillance until disease recurs. This leads to four branches: true positive (**TP**: the biomarker test is positive and RD is present), false positive (**FP**: biomarker test is positive but RD is not present), false negative (**FN**: the biomarker is negative but RD is present), and true negative (**TN**: the biomarker correctly identifies ypCR). Patients are sorted into each of these four branches according to the true probability of RD based on clinical trial outcomes of neoadjuvant therapy followed by RC^14,34–36^ and the sensitivity and specificity of the SaRCA biomarker test. Each of the five branches of the decision tree (RC for all, TP, FP, FN, and TN) leads to a MM, and the outcomes are admixed proportionally based on the sensitivity and specificity of the SaRCA test.

MMs for TP, FP, and FN have similar structure. Patients in the simulated cohort testing positively (TP and FP groups) undergo RC, with its attendant mortality, probability of complication, costs, survival, and QoL. Following surgery, the surviving cohort is established in a “no evidence of disease” (**NED**) state, whether or not residual disease is present. Note that all TP patients have RD; all FP patients achieved ypCR; and for the RC decision, the probability of RD is a variable that depends on the efficacy of NAC. Patients stay in the NED state until progressive disease (**PD**) is detected, death, or the end of the Markov simulation (20 years). Cycle length is six months, and, at each cycle, appropriate clinical visits and procedures are undertaken.

In the FN model, simulated patients have subclinical RD - a biomarker test indicates ypCR. For these patients, RC is delayed one cycle (i.e., six months), based approximately on outcomes of trials of RC avoidance, where some patients entering active surveillance protocols presented with prompt ‘recurrence’ that we surmise was subclinical disease^37,38^. From that point, the model proceeds as the TP case, with the exception that the probability of PD is increased by a hazard rate attributable to the delay^39^.

The TN model has more possible states than those of the other four. Patients with ypCR can stay in NED state, can have local superficial recurrence (**LSR**) which is treated with transurethral resection of bladder tumor (**TURBT**), locally advanced recurrence (**LAR**) treated with RC, or transition directly to PD or Death. If treated, they return to the NED state. In these situations the probability of progression is increased.

Data for the probability of RD, sensitivity and specificity of various candidate biomarkers, and the parameters of the MMs were taken from recent observational studies and clinical trials and summarized in **Supplemental Tables 1-4**.

## Results

The primary analysis was performed as above, based on outcomes with cisplatin-based NAC and published data to inform choice and chance trees and costs. A second analysis was performed to evaluate how emerging systemic therapies influence the value of testing. This analysis incorporated the EV-303/KEYNOTE-905 trial^34^; modeling the high post-treatment ypCR rate (57.1%) in cisplatin-ineligible patients receiving enfortumab vedotin + pembrolizumab. At the time of submission, EVP was approved by the FDA only for cisplatin-ineligible patients, but it is expected to become the new standard of care (**SoC**) for all MIBC patients who are RC candidates. All testing modalities are compared to a “no-test” strategy (i.e. the current SoC) and a perfect test which identifies residual disease with 100% sensitivity and specificity. Outcomes for RC avoidance are estimated from the RETAIN^38^, RETAIN-2^40^, and HCRN GU GU16-257^37^ trials, which used a combination of clinical factors to assign patients to therapy arms. We assumed that early ‘recurrences’ (those occurring within 6 months of entrance into active surveillance) were likely subclinical tumors missed upon clinical examination due to current limitations of residual disease detection tools and these cases were considered FN for the purpose of the MM. Costs, utilities, oncologic outcomes, and biomarker performance that animate the model are described in **Supplemental Tables 1-4**. Results are described below and fully documented in **Table 1**.

**Table 1:** MM outcomes when applying each SaRCA test to a cohort of RC candidates. Change in cost, life expectancy, and quality of life are expressed in comparison to a cystectomy-for-all scenario. All conditions except for guided biopsy result in decreased overall costs per patient. All conditions result in increased life expectancy and QoL. LE: life expectancy; QALY: quality-adjusted life years.

| Condition |  |  |  |  | Neoadjuvant MVAC |  |  |  | Neoadjuvant EVP |  |  |  |  |
| --- | --- | --- | --- | --- | --- | --- | --- | --- | --- | --- | --- | --- | --- |
|  | Cost of test | Sensitivity | Specificity |  | Baseline ΔCost | Baseline ΔLE | Baseline ΔQALY | Break-even cost of test |  | Baseline ΔCost | Baseline ΔLE | Baseline ΔQALY | Break-even cost of test |
| perfect test | \$2,900 | 100% | 100% | | -\$8,124 | +0.189 | +0.206 | \$11,024 | | -\$15,290 | +0.321 | +0.356 | \$18,190 |
| NTACT | \$2,900 | 91% | 50% | | -\$2,271 | +0.093 | +0.101 | \$5,171 | | -\$5,926 | +0.159 | +0.176 | \$8,830 |
| WashU Urine test | \$2,900 | 81% | 81% | | -\$5,310 | | +0.162 | \$8,210 | | -\$11,265 | | +0.284 | \$14,160 |
| ctDNA | \$2,900 | 59% | 100% | | -\$6,571 | | +0.196 | \$9,471 | | -\$14,062 | | +0.348 | \$16,960 |
| MRI | \$379 | 54% | 84% | | -\$7,139 | +0.152 | +0.162 | \$7,518 | | -\$13,523 | +0.263 | +0.290 | \$13,900 |
| Guided Bx | \$16,200 | 64% | 94% | | +\$7,201 | +0.172 | +0.185 | \$8,999 | | +\$179 | +0.297 | +0.327 | \$16,030 |

### Value of Testing with Neoadjuvant Cisplatin-Based Chemotherapy Regimen

We model a perfect test (100% sensitivity and specificity) to frame the maximal potential benefit that might be gleaned from a high performing biomarker. This perfect test yielded a cost savings of $8,124, a survival benefit of 2.3 months, and quality of life years improvement of 2.5 months compared to the current SoC featuring cisplatin-based chemotherapy and no option for RC avoidance. Despite moderate performance characteristics, each of the biomarker tests, including NTACT, WashU Urine test, and ctDNA as well as MRI demonstrated benefit across domains as compared to “no-test” strategy. SEE yielded more improved life expectancy and quality of life but was too expensive to be cost effective.

### Quality of Life

All testing strategies improved average quality of life scores with meaningful impact on overall QALY. ctDNA and SEE yielded the greatest improvement with 0.196 and 0.172 additional QALY (2.4 months, 2.2 months, respectively). These benefits derive from exemption from urinary diversion.

### Survival

Somewhat unexpectedly, there was a small survival benefit across testing modalities. The greatest benefit in average life expectancy was for guided biopsy at 0.172 years (2.1 months). The smallest benefit was for NTACT at 1.1 months. We believe this primarily stems from the high perioperative mortality rate associated with RC. We assumed that the fraction of the perioperative mortalities was proportionally distributed among patients who achieved pCR. RC avoidance in patients who would have otherwise achieved pCR is how surgical avoidance in some increases life expectancy for the whole pool of eligible patients on average.

### Healthcare Costs

Most testing strategies were cost-effective compared to not testing, with the greatest total healthcare expenditure reduction per patient of $7,139 for MRI and $6,571 for ctDNA. Only SEE yielded an increased cost compared to not testing at $7,201 due to the high cost of procedures under anesthesia and associated charges.

A subsequent break-even calculation was performed to determine the maximum cost at which each test yields a potential cost benefit. The break even cost for SEE was $8,999 (requiring a >$7,000 cost reduction).

### Value of Testing with Neoadjuvant Antibody Drug Conjugate and Immunotherapeutic Regimen

As expected, the benefit across domains was more pronounced with EVP due to the higher ypCR rate of this more active regimen. The perfect test model yielded an average cost savings of $15,290, a survival benefit of 3.9 months and additional QALY improvement of 4.2 months.

In the testing cohorts, EVP consistently demonstrates benefits to testing all versus not testing in the domains of healthcare costs, survival, and quality of life. Average survival benefit for testing all patients ranged from 3.6 to 1.9 months for SEE and NTACT, respectively. Calculated QALY benefit ranged from 4.2 to 2.1 months for WashU urine test and NTACT, respectively. Cost savings was noted in most modeled scenarios, with the largest cost savings for the WashU Urine test at $14,062. Guided biopsy had a calculated break even cost of $16,030, yielding a net additional cost of $179.

### Effect of ypCR rate on cost effectiveness

Considering how positively EVP affects the analyzed domains, therapies of the future that might produce even higher ypCR rates were considered. The change in costs and qoL were calculated against the likelihood of achieving ypCR, while holding the sensitivity and specificity of the test constant. For the case of the NTACT test, SaRCA is cost effective until the residual disease rate after neoadjuvant therapy is about 75%, regardless of whether the salvage systemic treatment is EVP or chemoimmunotherapy, although there is more potential for cost savings when the salvage systemic therapy is chemoimmunotherapy and residual disease rates are low (**Table 2**).

**Table 2:** The change in cost and QoL compared to a cystectomy-for-all approach was calculated under different pathologic residual disease rates using NTACT as a SaRCA test. Incremental cost effectiveness ratios were calculated when costs were increased compared to the cystectomy-for-all approach.

|  | Neoadjuvant MVAC |  | Neoadjuvant EVP |  |  |  |  |
| --- | --- | --- | --- | --- | --- | --- | --- |
| pRD rate | Baseline ΔCost | Baseline ΔQALY | Baseline ΔCost | Baseline ΔQALY |  | ICER value |  |
| 0 | -\$11,605 | 0.271 | -\$13,057 | 0.312 | Testing favored | | |
| 0.25 | -\$7,841 | 0.202 | -\$8,911 | 0.233 | Testing favored | | |
| 0.5 | -\$4,078 | 0.134 | -\$4,765 | 0.154 | Testing favored | | |
| 0.75 | -\$314 | 0.065 | -\$619 | 0.075 | Testing favored | MVAC | EVP |
| 0.8 | +\$439 | 0.051 | +\$210 | 0.059 | RC favored | \$8,608 | \$3,559 |
| 0.85 | +\$1,191 | 0.038 | +\$1,039 | 0.043 | RC favored | \$31,342 | \$24,163 |
| 0.9 | +\$1,944 | 0.024 | +\$1,869 | 0.027 | RC favored | \$81,000 | \$69,222 |
| 0.95 | +\$2,697 | 0.010 | +\$2,698 | 0.012 | RC favored | \$269,700 | \$224,833 |
| 1 | +\$3,450 | -0.003 | +\$3,527 | -0.004 | RC favored | | |

## Discussion

There is great enthusiasm to avoid RC, and several approaches to prospectively identify patients achieving ypCR have been reported. For instance, tissue biopsy prior to RC identifies responders but unfortunately also fails to identify many patients who have residual disease in a prospective study^41^. Retrospective analysis of RC avoidance cohorts illustrates the likely consequence of understaging occult disease^15–17,42,43^. Tissue biomarkers such as those identified through targeted sequencing approaches show strong association with responder status^44,45^ but also may have limitations^46^. Nevertheless, several groups are working to solve this problem.

In our study, we found that most approaches to responder status assignment are viable. It seems that the specificity rate of the test drives cost savings (negative tests drive SaRCA) while sensitivity drive safety (patients with residual disease are allocated to RC).

Technical and practical issues require consideration prior to implementation of a SaRCA test. First, a SaRCA test would need to be developed and optimized. Secondly, stakeholders such as patients, physicians, insurance companies, and regulatory bodies would need to determine when and how the test could be applied to result in maximum benefit. As currently practiced, treatment of MIBC uses a “definitive-therapy-for-all approach” when feasible. However, in some cases, ‘definitive therapy’ could include neoadjuvant therapy alone, and possibly even TURBT alone. Although we illustrate the full capacity to reduce cost and improve morbidity and survival to which providers can aspire, no test will result in residual disease for every RC patient and long-term bladder-intact and metastasis-free survival. Modeling the impact of the test on clinical outcomes would help stakeholders make informed decisions about how and when to apply a SaRCA test.

We performed an analysis to determine performance conditions under which a SaRCA test, would be cost effective and how it would impact QALY and survival regardless of which decision making tool is used. We found that the biomarker can be cost effective and improve QoL and survival performing under conditions that seem very achievable. The key findings from the study are three-fold.

Firstly, biomarkers for SaRCA are cost effective. Improved tests and improved therapies that yield higher ypCR rates will result in improved cost effectiveness. Interestingly, as ypCR rates improve, a SaRCA test won’t even need to be that accurate because the pretest probability of response is so high – this engenders forgiveness for erroneous tests. If applied on a large scale, a SaRCa test could result in significant cost savings. These types of data might inform market pricing of such a test as well. This analysis is limited by the assumptions we made based on the published literature and would be impacted by such factors as local cost of RC, complication rates and associated costs, recurrence rates and treatment of local or distant recurrence, and intensity of surveillance after SaRCA. As there is no standardized approach to surveillance after SaRCA, a standard similar to that used for surveillance of NMIBC was employed.

Secondly and perhaps surprisingly, an effective SaRCA test realizes a small overall survival advantage. This increase is likely related to SaRCA in patients who would have achieved ypCR and would have otherwise had a long expected overall survival if they did not undergo RC and did not have residual disease. The survival benefit of SaRCA test use would increase or decrease with an increase or decrease, respectively, in estimated perioperative mortality rates. This analysis assumes that perioperative mortality rate is randomly distributed among responders and nonresponders, but this assumption may not be true. Studies to inform that assumption are lacking.

Third, and expectedly, the use of a SaRCA test is associated with improved QoL. QALY are increased in all sensitivity and specificity scenarios tested compared to a ‘no testing’ strategy. This is likely reflective of the presumed decrement in QoL after RC and the anticipated improvement in QoL under SaRCA conditions. Literature on which to base these assumptions is sparse compared to literature describing RC-associated costs and survival, so this portion of the model is the most likely to change with better data to inform the MM.

We did not model how a SaRCA test would impact costs, survival, and QoL for patients undergoing trimodal therapy with TURBT, neoadjuvant chemotherapy, and consolidative chemoradiation. It is not clear which portion of trimodal therapy could be omitted based on a biomarker of response. Perhaps radiation (the most expensive modality of trimodal therapy) could be omitted, however, this has not been studied and there is no discernible effort to omit radiation that we could identify. Further, trimodal therapy is associated with a different QoL profile. Although many patients undergoing trimodal therapy have long bladder intact survival, those who do have a different set of QoL issues relating to complications from radiation, such as bowel or bladder dysfunction, post radiation strictures of the genitourinary or gastrointestinal tract, and management of hematuria and hematochezia. These morbidities could be reduced with the omission of pelvic radiation. Further, patients undergoing salvage RC after trimodal therapy have a different complication profile as they are much more likely to undergo urinary and stool diversion than patients undergoing RC in the absence of prior radiation. Mortality rates related to the trimodal therapy are expected to be much lower. How a SaRCA test would perform under these conditions and for what purpose a test could be developed is unknown.

In summary, we show that a SaRCA test is expected to be cost effective, improve QoL, and overall survival in patients with MIBC undergoing neoadjuvant chemotherapy and RC.

Development of SaRCA algorithms would be a benefit to patients, caregivers, and the healthcare system.

## Supporting information

Supplemental Table 1_Cost Table

Supplemental Table 2_Survival Tables

Supplemental Table 3_QoL Variables Table

Supplemental Table 4_SS Variables Table

## Data Availability

All data in the present work are contained in the manuscript and supplemental tables.

## Acknowledgments and funding

This study was supported by Fox Chase Cancer Center support grant P30 CA006927 and grant CA260369 (PHA). PHA has patent pending on urine biomarker testing for residual disease.

---

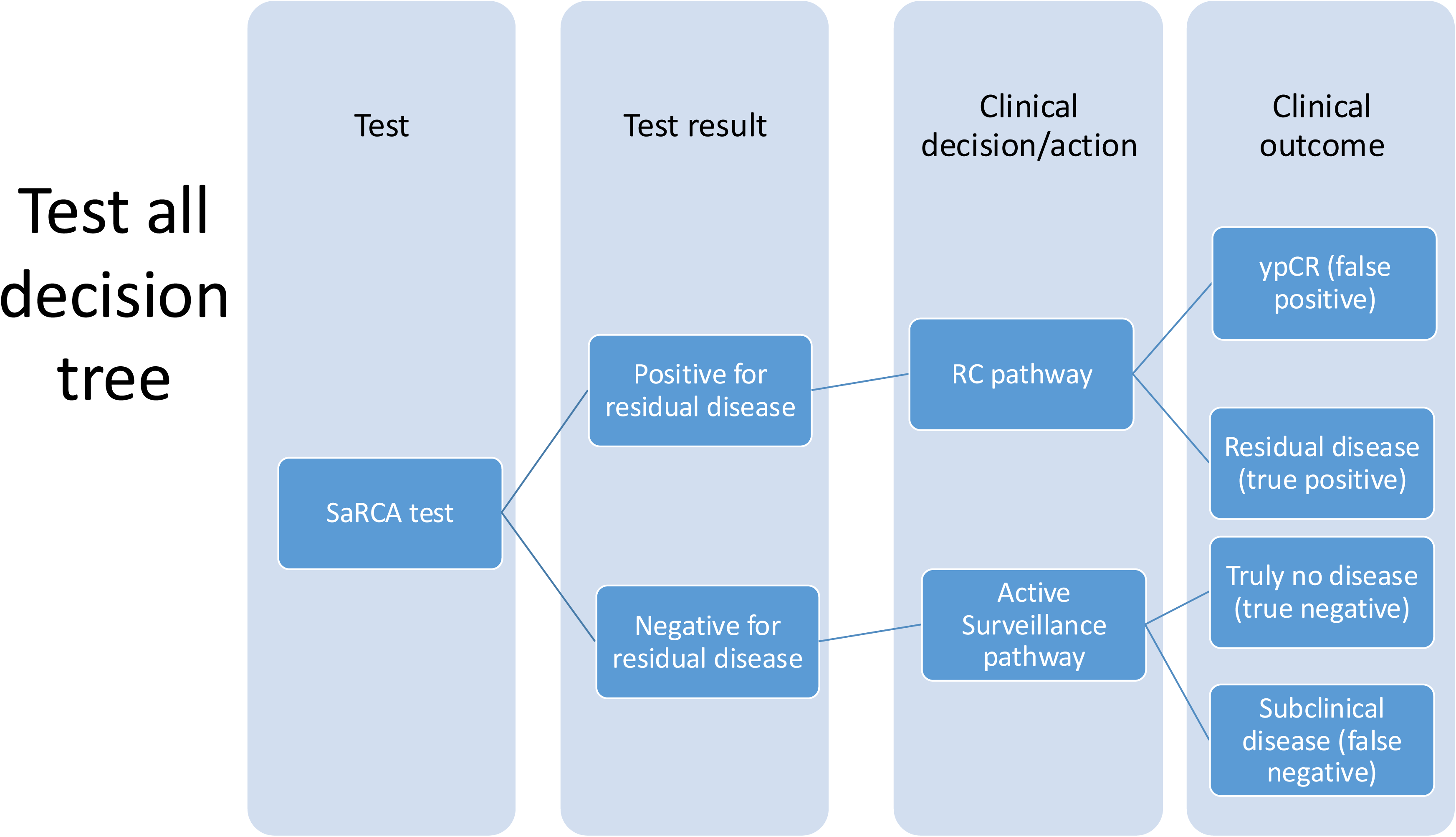

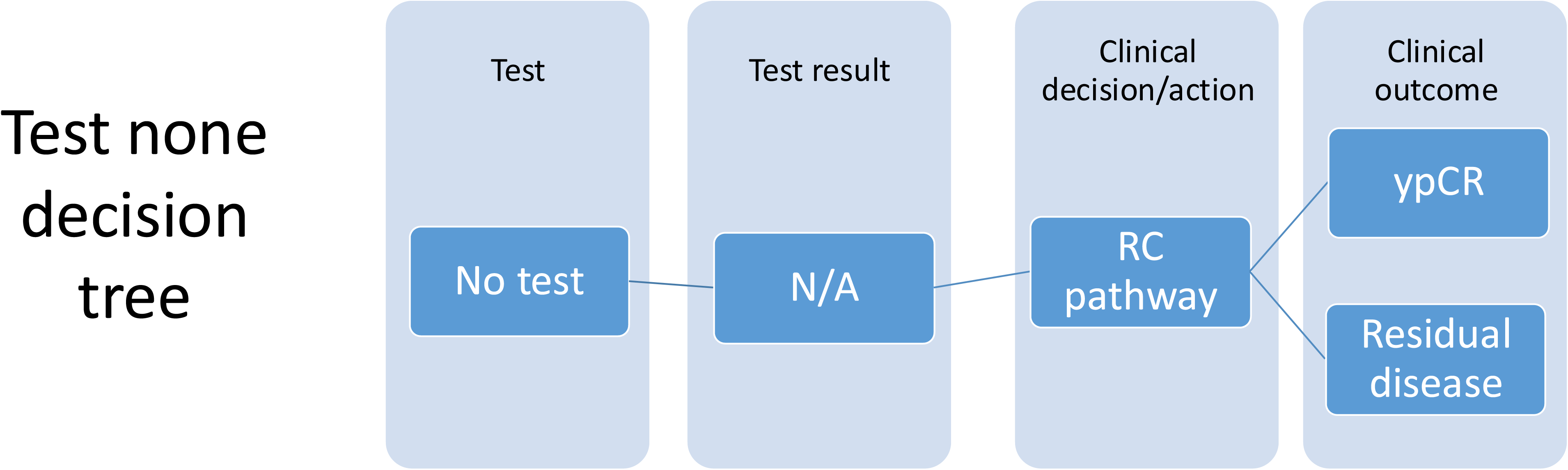

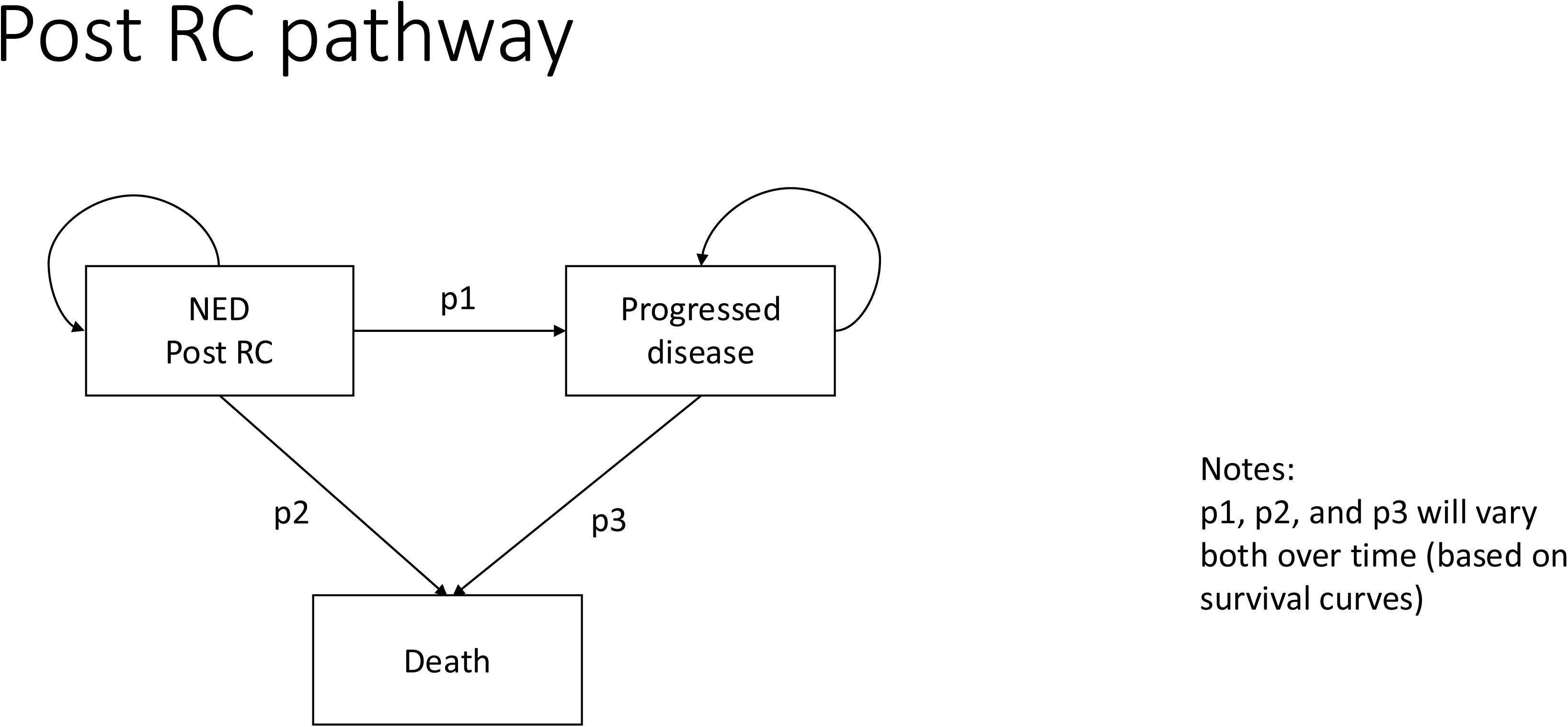

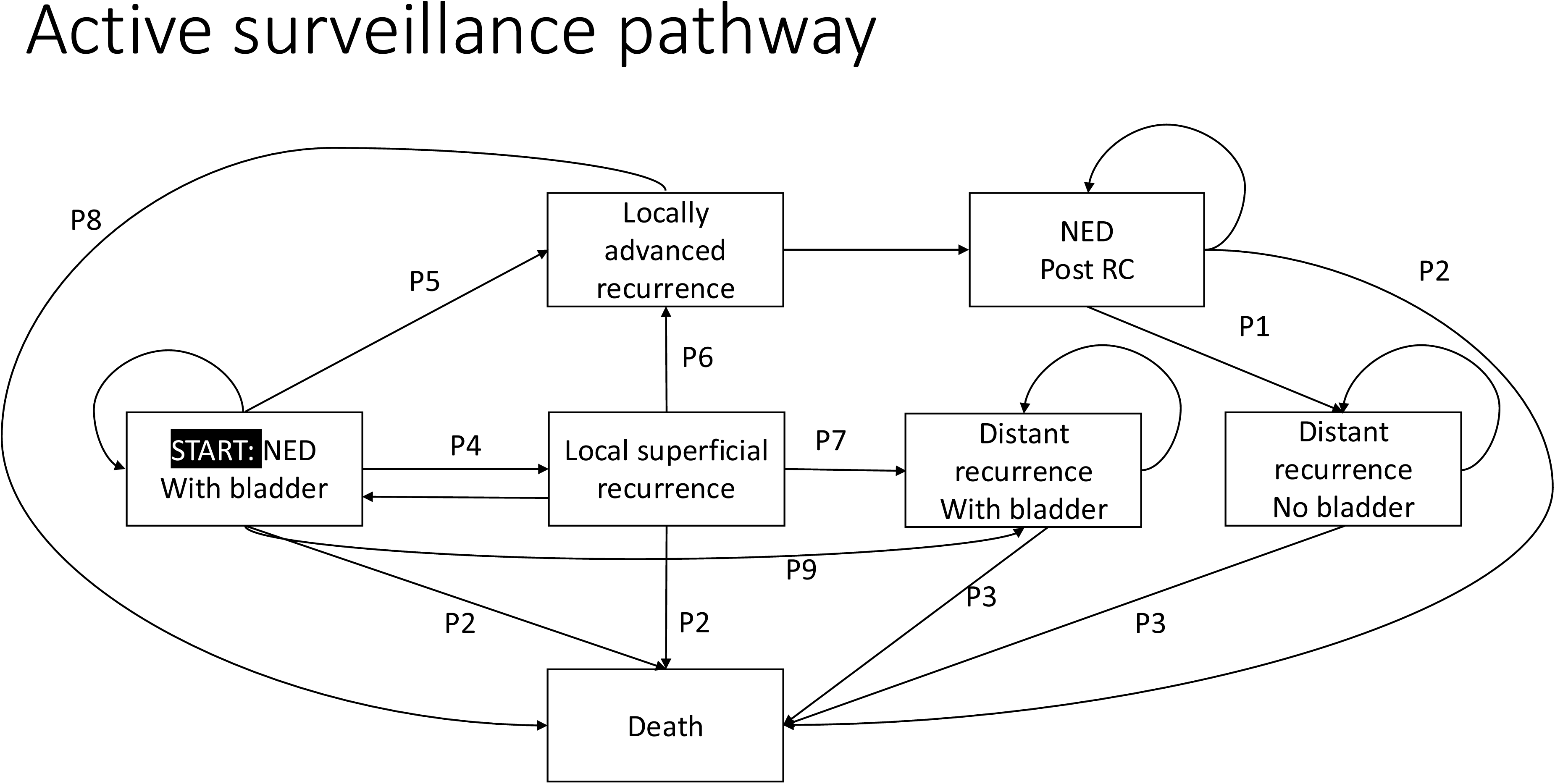

## Notes

### Competing Interest Statement

The authors have declared no competing interest.

