## Supplemental Table 1_Cost Table for "Modeling Biomarker-Guided Avoidance of Radical Cystectomy: Costs and Outcomes"

**Supplemental Table 1: Model Inputs for costs associated with each clinical scenario.**

| **Model Node** | **Parameter** | **Cycles** | **Procedure Cost** | **Reference** |
| --- | --- | --- | --- | --- |
| **SaRCA test negative, residual disease present (False Negative), RC avoidance followed by 6-month delayed RC** | NED post RC | 1-4 | Surveillance:  Physician visit (99213): $168.75  Cytology (88147): $63.25  chest CT (71260) + CT abd/pelv (74178) $446 | Medicare table |
|  | Progressive Disease (metastasis post RC) | 1-4 | PET CT (78814): $1,604 | Medicare table |
|  |  |  | Percutaneous lung biopsy (32408): $1,113 | Medicare table |
|  |  |  | EVP $113,691 per cycle (after cisplatin-based neoadjuvant therapy)  Gemcitabine + platinum-based chemotherapy + anti PD1 immunotherapy: $ 41,313 (after neoadjuvant EVP) | Chiddarwar et al^1^ ($752,637 total cost ÷ 3.31 years ÷ 2 cycles per year)  Meng et al^2^ (223,091 total cost ÷ 2.7 years ÷ 2 cycles per year) |
|  |  |  | Surveillance:  Physician visit (99213): $168.75  Cytology (88147): $63.25  chest CT (71260) + CT abd/pelv (74178) $446 | Medicare table |
|  | Death | All | Death | NA |

| **SaRCA test positive, residual disease present (true positive) undergoes immediate RC** | NED post RC | 1-4 | Surveillance:  Physician visit (99213): $168.75  Cytology (88147): $63.25  chest CT (71260) + CT abd/pelv (74178) $446 | Medicare table |
| --- | --- | --- | --- | --- |
|  | Progressive Disease (metastasis post RC) | 1-4 | PET CT (78814): $1,604 | Medicare table. |
|  |  |  | Percutaneous lung biopsy (32408): $1,113 | Medicare table. |
|  |  |  | EVP $113,691 per cycle (after cisplatin-based neoadjuvant therapy)  Gemcitabine + platinum-based chemotherapy + anti PD1 immunotherapy: $ 41,313 (after neoadjuvant EVP) | Chiddarwar et al^1^ ($752,637 total cost ÷ 3.31 years ÷ 2 cycles per year)  Meng et al^2^ (223,091 total cost ÷ 2.7 years ÷ 2 cycles per year) |
|  | Death | All | Death | NA |
| **SaRCA test positive, no residual disease present (false positive) undergoes immediate RC** | NED post RC | 1-4 | Surveillance:  Physician visit (99213): $168.75  Cytology (88147): $63.25  chest CT (71260) + CT abd/pelv (74178) $446 | Medicare table |
|  | Death | All | Death | NA |

| **SaRCA test negative, no residual disease present (true negative), RC avoidance** | NED with Bladder | 1-4 | Surveillance:  Physician visit (99213): $168.75  Cytology (88147): $63.25  chest CT (71260) + CT abd/pelv (74178) $446 | Medicare table |
| --- | --- | --- | --- | --- |
|  | Local Advanced Recurrence |  | TURBT: $16,163  RC: $39,720 |  |
|  | Local Superficial Recurrence |  | TURBT: $16,163 |  |
|  |  |  | Intravesical BCG: $1,085 | Scilipoti et al^3^ |
|  |  |  | Intravesical Mitomycin:$296 - $1,262 | Scilipoti et al^3^ |
|  |  |  | Surveillance:  Physician visit (99213): $168.75  Cytology (88147): $63.25  chest CT (71260) + CT abd/pelv (74178): $446  Cystoscopy (52000): $250 | Medicare table |
|  | Distal Recurrence With Bladder |  | PET CT (78814): $1,604 | Medicare table |
|  |  |  | Percutaneous lung biopsy (32408): $1,113 | Medicare table |
|  |  |  | EVP $113,691 per cycle (after cisplatin-based neoadjuvant therapy)  Gemcitabine + platinum-based chemotherapy + anti PD1 immunotherapy: $ 41,313 (after neoadjuvant EVP) | Chiddarwar et al^1^ ($752,637 total cost ÷ 3.31 years ÷ 2 cycles per year)  Meng et al^2^ ($223,091 total cost ÷ 2.7 years ÷ 2 cycles per year) |
| **No testing**  **All Comers undergo RC** | NED post RC | 1-4 | Surveillance:  Physician visit (99213): $168.75  Cytology (88147): $63.25  chest CT (71260) + CT abd/pelv (74178): $446  Cystoscopy (52000): $250 | Medicare table |
|  | Death | All | Death | NA |

EV: enfortumab vedotin; NA: not applicable; NED: no evidence of disease; RC: radical cystectomy
