## Supplemental Table 2_Survival Tables for "Modeling Biomarker-Guided Avoidance of Radical Cystectomy: Costs and Outcomes"

**Supplementary Table 2: Recurrence-free and overall survival outcomes in different clinical scenarios.**

| Model Node | Parameter | Description | Numeric Input | Reference |
|---|---|---|---|---|
| **SaRCA test negative, residual disease present (False Negative), RC avoidance followed by 6-month delayed RC** | pA1 | Probability of developing progressed disease within 6 months post radical cystectomy | HR per day: 1.001, 95% CI (1.000-1.002) | Kulkarni et al[1] |
| | pA2 | Mortality starting at age 70 from lifetables | | Period Life Table, 2002, as used in the 2025 Trustees Report. Social Security Administration. https://www.ssa.gov/oact/STATS/table4c6.html |
| | pA3 | Median overall survival with metastatic disease (treatment with EVP) | 31.5 months | Powles et al[2] |
| **SaRCA test positive, residual disease present (true positive) undergoes immediate RC** | pB1 | 10-year disease-specific survival based on tumor grade and stage (pT2a/b pN0) | 66.8% | Hautmann et al[3] |
| | | 10-year disease-specific survival based on tumor grade and stage (pT3a/b pN0) | 59.7% | |
| | | 10-year disease-specific survival based on tumor grade and stage (pT4a/b pN0) | 36.6% | |
| | | 10-year disease-specific survival based on tumor grade and stage (pTany, pN+) | 16.7% | |
| | | 5-year recurrence free | 45% | Bhindi B et al[4] |
| | pB2 | Mortality starting at age 70 from lifetables | | Period Life Table, 2002, as used in the 2025 Trustees Report. Social Security Administration. https://www.ssa.gov/oact/STATS/table4c6.html |
| | pB3 | Median overall survival with metastatic disease | 31.5 months | Powles et al[2] |

| | | | | |
|---|---|---|---|---|
| **SaRCA test positive, no residual disease present (false positive) undergoes immediate RC** | pC1 | 10-year disease-specific survival based on tumor grade and stage (pT0/a/is/1, pN0) | 90.5% | Hautmann et al[3] |
| | | 5-year RFS for tumor ypT0N0 | 90.0% | Bhindi B et al[4] |
| | pC2 | Mortality starting at age 70 from lifetables | | Period Life Table, 2002, as used in the 2025 Trustees Report. Social Security Administration. https://www.ssa.gov/oact/STATS/table4c6.html |
| | pC3 | Median overall survival with metastatic disease | 31.5 months | Powles et al[2] |
| **SaRCA test negative, no residual disease present (true negative), RC avoidance** | P1 | 10-year disease-specific survival based on tumor grade and stage (pT2a/b pN0) | 66.8% | Hautmann et al[3] |
| | | 10-year disease-specific survival based on tumor grade and stage (pT3a/b pN0) | 59.7% | Hautmann et al[3]<br>Bhindi B et al[4] |
| | | 10-year disease-specific survival based on tumor grade and stage (pT4a/b pN0) | 36.6% | |
| | | 10-year disease-specific survival based on tumor grade and stage (pTall pN+) | 16.7% | |
| | | 5-year RFS | 45% | |
| | P2 | Mortality starting at age 70 from lifetables | | Period Life Table, 2002, as used in the 2025 Trustees Report. Social Security Administration. https://www.ssa.gov/oact/STATS/table4c6.html |
| | P3 | Median overall survival with metastatic disease | 31.5 months | Powles et al[2] |
| | P4 | Local superficial at 33 months | 1.63% per cycle | Galsky et al[5] |
| | | 3 years (exponential) | 9.375% | Galsky et al[5] |
| | P5 | Local advanced at 18 months | 12.5% | Galsky et al[5] |

| | | | | |
|---|---|---|---|---|
| | P6 | Locally advanced disease after superficial recurrence | 2% per cycle | Galsky et al[5] |
| | P7 | Distant recurrence after superficial-only recurrence | 1% lifetime | |
| | P8 | Mortality from locally advanced recurrence without RC | 3.7% (1 month)<br>2% (3 months)<br>3% (3 months)<br>3% (3 months) | International Collaboration of Trialists[6]<br>Catto et al[7]<br>Novara et a[8]<br>Parekh et al[9] |
| | P9 | Metastatic recurrence without local recurrence at 55 months | 4% lifetime | Geynisman[10] |
| | | Per 6-month cycle after 3 years | 1% | |
| **Scenario E: All Comers, got RC** | pE1 | 5-year survival (including death as an event) | 52.8% | Grossman et al[11] |
| | pE2 | Mortality starting at age 70 from lifetime tables | | Period Life Table, 2002, as used in the 2025 Trustees Report. Social Security Administration. https://www.ssa.gov/oact/STATS/table4c6.html |
| | pE3 | Mortality overall survival with metastatic disease (treatment with EVP) | 31.5 months | Powles et al[2] |

NA: not applicable; RC: radical cystectomy; RD: residual disease; RFS: recurrence free survival
