## Supplemental Table 3_QoL Variables Table for "Modeling Biomarker-Guided Avoidance of Radical Cystectomy: Costs and Outcomes"

**Supplemental Table 3:** **Quality of life of bladder cancer patients with differing stages of disease.**

| **Scenario** | **Condition** | **QoL**  **(Global Mean Score)** | **Reference** |
| --- | --- | --- | --- |
| **NED with Bladder** | TURBT-only | 70.0 | Catto et al^1^ |
|  | TURBT & BCG/MMC | 71.9 |  |
|  | Radical Radiotherapy treatment | 68.6 |  |
|  | TMT | 80 | Mak et al^2^ |
| **Local Advanced Recurrence** | Baseline | 73 | Taarnhøj et al^3^ |
|  | After 4 cycles of treatment | 72 |  |
| **Local Superficial Recurrence** | 6 weeks after diagnosis of non-muscle invasive BCa | 76 | Beeren et al^4^ |
|  | 3 months after diagnosis of non-muscle invasive BCa | 75.8 |  |
|  | 15 months after diagnosis of non-muscle invasive BCa | 77.4 |  |
|  | 51 months after diagnosis of non-muscle invasive BCa | 78.6 |  |
| **Distal Recurrence with Bladder** | Metastatic BCa (baseline) | 54 | Taarnhøj et al^3^ |
|  | Metastatic BCa (After 3 cycles of treatment) | 69 |  |
| **Progressive Disease** | Metastatic BCa | 64.3 | Smith et al^5^ |

BCa: bladder cancer; MMC: intravesical mitomycin C; NED: no evidence of disease; QoL: quality of life; RC: radical cystectomy; TMT: Bladder-sparing trimodality therapy

QoL measurement: European Organization of Research and Treatment of Cancer (EORTC) QLQ-C30 questionnaire.

Note: some values were adjusted slightly to account for differences in study outcomes in order to make cross-study comparisons consistent.
