## Supplemental Table 4_SS Variables Table for "Modeling Biomarker-Guided Avoidance of Radical Cystectomy: Costs and Outcomes"

**Supplemental Table 4: Sensitivity and specificity of chosen SaRCA tests**

|  | **Sensitivity** | **Specificity** | **Reference** |
| --- | --- | --- | --- |
| perfect test | 100% | 100% |  |
| NTACT | 91% | 50% | Satyal et al^1^ |
| WashU Urine test | 81% | 81% | Chauhan at el^2^ |
| ctDNA | 59% | 100% | Ben-David et al^3^ |
| MRI | 54% | 84% | Necchi et al^4^ |
| SEE | 64% | 94% | Zibelman et al^5^ |
